# EARLY LENZILUMAB TREATMENT OF COVID-19 PATIENTS USING C-REACTIVE PROTEIN AS A BIOMARKER IMPROVES EFFICACY: RESULTS FROM THE PHASE 3 ‘LIVE-AIR’ TRIAL

**DOI:** 10.1101/2021.12.30.21267140

**Authors:** Zelalem Temesgen, Colleen F. Kelley, Franklin Cerasoli, Adrian Kilcoyne, Dale Chappell, Cameron Durrant, Omar Ahmed, Gabrielle Chappell, Victoria M. Catterson, Christopher Polk, Andrew D. Badley, Vincent C. Marconi, the LIVE-AIR Study Group

**Affiliations:** Mayo Clinic, Division of Infectious Disease, Rochester, MN; Division of Infectious Diseases, Emory University School of Medicine, Grady Memorial Hospital, Atlanta, GA; RxMedical Dynamics, New York, NY; Humanigen, Inc., Burlingame, CA; BioSymetrics, Inc., New York, NY; Atrium Health, Charlotte, NC; Mayo Clinic, Division of Infectious Disease and Department of Molecular Medicine, Rochester, MN; Division of Infectious Diseases, Emory University School of Medicine, Rollins School of Public Health, and Emory Vaccine Center, Atlanta, GA

**Author notes:** **Corresponding author address** Zelalem Temesgen, MD, Mayo Clinic, 200 First St. SW Rochester, MN 55901.

**Keywords:** Lenzilumab, GM-CSF, COVID-19, hyperinflammation, SARS-CoV-2, LIVE-AIR, CRP

## Abstract

**Objective:** The LIVE-AIR trial demonstrated that the anti-GM-CSF monoclonal antibody, lenzilumab improved the likelihood of survival without invasive mechanical ventilation (SWOV) in COVID-19 patients; with greatest effect in those with baseline CRP below the median baseline value of 79 mg/L. Similar to GM-CSF, C-reactive protein (CRP) levels are correlated with COVID-19 severity. This current analysis assessed the utility of baseline CRP levels to guide treatment with lenzilumab.

**Design:** LIVE-AIR was a phase 3, double-blind, placebo-controlled trial. Participants were randomized 1:1 and stratified according to age and disease severity, to receive lenzilumab or placebo on Day 0, were followed through Day 28.

**Setting:** Secondary and tertiary care hospitals in the US and Brazil.

**Participants:** 520 hospitalized COVID-19 participants with SpO2≤ 94% on room air or required supplemental oxygen but not invasive mechanical ventilation were included.

**Interventions:** Lenzilumab (1800mg; divided as 3 doses, q8h) or placebo infusion alongside standard treatments including corticosteroids and remdesivir.

**Main outcome measures:** A multi-variate logistic regression analysis assessed key baseline risk factors for progression to IMV or death. The primary endpoint, SWOV, and key secondary endpoints were analyzed according to baseline CRP levels in all participants with CRP values.

**Results:** The multi-variate analysis demonstrated that elevated baseline plasma CRP was the most predictive feature for progression to IMV or death. SWOV was achieved in 152 (90%; 95%CI: 85to 94) lenzilumab and 183 (79%; 72 to 84) placebo participants with baseline CRP<150 mg/L and its likelihood was greater with lenzilumab than placebo (HR: 2.54; 95%CI, 1.46 to 4.41; p=0.0009) but not in participants with CRP≥150 mg/L at baseline. CRP as a covariate in the overall analysis demonstrated a statistically significant interaction with lenzilumab treatment (p=0.044). Grade ≥ 3 adverse events in participants with baseline CRP<150 mg/L were reported in 18% and 28% in lenzilumab or placebo, respectively. No treatment-emergent serious adverse events were attributable to lenzilumab.

**Conclusion:** These finding suggest that COVID-19 participants with low baseline CRP levels achieve the greatest clinical benefit from lenzilumab and that baseline CRP levels may be a useful biomarker to guide therapeutic intervention.

**Trial Registration:** ClinicalTrials.gov NCT04351152

**WHAT IS ALREADY KNOWN ON THIS TOPIC:** GM-CSF is one of the early upstream mediators and orchestrators of the hyperinflammatory immune response following SARS-CoV-2 infection. Baseline levels of GM-CSF and CRP have each been shown to correlate with COVID-19 disease progression. Increases in CRP are driven by elevations of IL-6 during the hyperinflammatory response following SARS-CoV-2 infection. In the phase 3, randomized, double-blind, placebo-controlled LIVE-AIR study, GM-CSF neutralization with lenzilumab significantly improved the likelihood of survival without invasive mechanical ventilation (SWOV, primary endpoint, also referred to as ventilator-free survival) vs. placebo (HR:1.54; 95% CI, 1.02 to 2.32; p=0.0403), which included standard supportive care including corticosteroids and remdesivir. No treatment-emergent serious adverse events attributable to lenzilumab have been reported to date.

**WHAT THIS STUDY ADDS:** A comprehensive analysis of LIVE -AIR CRP data provides evidence for the utility of baseline CRP to predict progression to IMV and death. Baseline CRP was identified to be the strongest predictor of SWOV in this study. Patients with baseline CRP<150 mg/L represented 78% of the study population and demonstrated the greatest clinical benefit with lenzilumab, including SWOV through day 28 (HR: 2.54; 95%CI; 1.46-4.41; nominal p=0.0009). A biomarker-driven approach using baseline CRP levels to guide therapeutic intervention may improve outcomes in those hospitalized with COVID-19. Participants with baseline CRP levels above 150 mg/L were described as experiencing COVID-19-associated hyperinflammation and were at risk of imminent escalation of respiratory support or death. Elevated baseline plasma CRP was the most predictive feature for progression to IMV or death (OR, 0.15; 95%CI, 0.07-0.29; nominal p<0.001). These findings suggest that baseline CRP may be a useful biomarker in determining which participants may be most successfully treated with lenzilumab.

## INTRODUCTION

A hyperinflammatory response, characterized by activation and trafficking of myeloid cells, increased secretion of downstream inflammatory chemokines (MCP-1, IL-8, IP-10), cytokines (IL-6, IL-1)^1^, and markers of systemic inflammation (CRP, D-dimer, ferritin), has been implicated in the morbidity and mortality due to COVID-19. ^1-4^ Granulocyte-macrophage-colony-stimulating factor (GM-CSF) is one of the early upstream mediators and orchestrators of this hyperinflammatory immune response. Increasing levels of circulating GM-CSF have been associated with progression and increasing severity of disease.^1^

Similar to GM-CSF, C-reactive protein (CRP) levels directly correlate with COVID-19 disease severity. ^1^ Increases in CRP are driven by elevations of IL-6 during the hyperinflammatory response following SARS-CoV-2 infection. ^5,6^ Baseline CRP levels predict subsequent oxygen supplementation requirements in hospitalized patients with COVID-19 patients from 85 mg/L for those on low-flow O_2,_; to 110 mg/L for those on high-flow O_2;_ and 205 mg/L for those on invasive mechanical ventilation (IMV). ^1^ Baseline CRP levels were also significantly higher in patients who had worsening organ failure (defined as an increase of sequential organ failure assessment [SOFA] score ≥ 1 point; compared to patients without worsening organ failure (mean CRP of 178 mg/L vs 100 mg/L, respectively, p<0.05).^7^ The risk of critical illness among hospitalized patients with CRP>200 mg/L was 2-fold greater compared with CRP between 15-100 mg/L (OR, 5.1; 95%CI: 2.8-9.2 vs 2.4; 95% CI: 1.4-4.0, respectively).^8^ The thirty-day risk of ICU admission or death progressively increased with CRP levels; 21.5% (95% CI: 18.1 to 24.9) in patients with baseline CRP levels of ≤ 99 mmol/L (99 mg/L) and 39.2% (95% CI: 35.6 to 43.0) in patients with baseline CRP levels of 100 to 400 mmol/L; 100 to 400 mg/L; p<0.001).^9^ Risk of 30-day mortality was similarly increased for patients with elevated CRP levels (p<0.001): normal CRP (7%; 0 to 15), CRP levels above normal but ≤ 99 mmol/L (18%; 15 to 21) and CRP of 100 to 400 mmol/L (29%; 5 to 32).^9^ Patients with CRP above 150 mg/L were described as experiencing COVID-19-associated hyperinflammation and were at risk of imminent escalation of respiratory support or death.^10^ The use of plasma CRP as a guide treatment is emerging. For example, the efficacy of corticosteroids in COVID-19 treatment has recently been associated with CRP levels. ^11^ and models are being developed in which CRP can be included for treatment guidance. ^12^

Lenzilumab, a GM-CSF neutralizing monoclonal antibody, has been shown in the LIVE-AIR phase 3 clinical trial to improve clinical outcomes in hypoxemic hospitalized patients with COVID-19.^13^ Lenzilumab was administered to COVID-19 patients with decreased oxygen saturation (SpO2≤ 94%) on room air, or who required supplemental oxygen but not yet on invasive mechanical ventilation (IMV) and within a median of 2 days after hospitalization. Lenzilumab improved the likelihood of survival without ventilation (SWOV, sometimes referred to as ventilator-free survival; HR:1.54; 95%CI, 1.02 to 2.32; p=0.0403) compared with placebo.^13^ A univariate sensitivity analyses of the primary endpoint for baseline factors that may influence the primary analysis demonstrated that baseline plasma CRP values below the median level of 79 mg/L were associated with a greater likelihood of achieving SWOV, relative to placebo (HR: 2.71; 95%CI: 1.23 to 6.00; nominal p=0·014) than in the overall population. ^13^

Given the above findings, we hypothesized that the earlier treatment of hyperinflammatory immune response with lenzilumab could be guided by clinical evaluation of CRP levels at presentation. CRP may be used as a practical and readily available biomarker in routine clinical practice^9,14^ that could predict which patients were suitable for “early” intervention with lenzilumab to prevent progression to IMV or death. Therefore, the objective of the following analyses of the LIVE-AIR trial was to demonstrate the utility of CRP as a prognostic biomarker to guide the treatment of COVID-19 with lenzilumab.

## METHODS

The LIVE-AIR trial design has been previously described in detail ^13^ and is briefly summarized here.

### Trial Design

LIVE-AIR is a randomized, double-blind, placebo-controlled, phase 3 trial (NCT04351152) and enrolled hospitalized participants with COVID-19 pneumonia. Eligibility criteria included age 18 years or older, virologically confirmed SARS-CoV-2, and pneumonia diagnosed by chest x-ray or computed tomography. Participants must have been hospitalized with a clinical ordinal score of 5 (SpO2 ≤ 94% on room air) or clinical ordinal score of 4 (supplemental oxygen in the form of low-flow oxygen) or clinical ordinal score 3 (high-flow oxygen, or non-invasive positive pressure ventilation) adapted from the NIH-sponsored Adaptive COVID-19 Treatment Trial (ACTT, NCT 04280705).^15^ Enrolled participants were randomized 1:1 to receive lenzilumab or matched placebo in addition to current standard treatments per institutional guidelines at each site. Three doses of lenzilumab (1800 mg total, divided into three doses) or placebo were administered 8 hours apart via a 1-hour IV infusion per dose. Participants were stratified by age (<65 or >65) and disease severity (severe vs. critical). The primary efficacy endpoint was SWOV by Day 28. For purposes of the survival analysis for the primary endpoint, an event was defined as mortality or the requirement for IMV. Secondary endpoints included time to recovery, the proportion of the composite of IMV (ordinal score 2), ECMO (ordinal score 2) or death (ordinal score 1); ventilator-free days; duration of ICU; mortality, and safety.

### Statistical Analysis

The primary endpoint was the difference between lenzilumab treatment and placebo treatment, in addition to standard treatments including remdesivir and dexamethasone, in SWOV through 28 days following randomization in the prespecified modified intent to treat population (mITT) who received at least one dose of investigational treatment under the documented supervision of the principal investigator or sub-investigator. This population was defined as the primary analysis and a Cox proportional hazard model (HR: lenzilumab relative to placebo) accounting for the stratification variables (i.e., age and disease severity) was used, supplemented by a display of K-M curves in each treatment group. The Cox proportional hazard model included the time to first event (death or IMV) as the dependent variable, (1=IMV use or death, 0=alive with no IMV use); treatment (covariate); and strata (covariates). Where data were non-proportional based on a Chi-squared test proposed by Grambsch and Therneau with a global p-value <0.05, a Cox proportional hazard model with weighted extension was used to correct for non-proportionality. Baseline CRP values were determined based on the screening value and if the participant did not have a screening value, then the day 1 value was used.

For each secondary endpoint, the proportion of participants that had the event was calculated by treatment group. An odds ratio was calculated for the composite endpoint of the first incident of IMV, ECMO, or death using logistic regression and including the baseline age group and disease category as covariates. For ventilator-free days and duration of ICU, the ANCOVA model of normality assumption was found to be clearly violated (e.g., p<0·05 for the Shapiro-Wilk test for normality), so a sensitivity analysis was conducted using an alternative non-parametric approach. A negative binomial regression model that was specified in the SAP was used, although the data did not conform to a Pascal distribution. Given that the data are not a Pascal distribution, a nonparametric stratified Wilcoxon test was performed using age strata and disease severity strata as stratification variables. Hazard ratios were calculated for each of time to death and time to recovery, separately, as described above. For time to recovery, deaths were censored at Day 28. Participants who were alive, yet did not recover, were right censored at the date of the last non-missing assessment of the 8-point clinical status ordinal scale on or prior to Day 28. All data reported herein are reported through Day 28. Loss to follow-up was approximately 2% in each arm with only 11 participants (5 and 6 in lenzilumab and placebo, respectively) in the mITT who had no vital status at Day 28. Of these 11 participants, 7 had recovered and were discharged and subsequently lost to follow-up. Four participants withdrew from the study prior to day 28 (2 lenzilumab and 2 placebo). Given the limited amount of missing data, the last observation carried forward method was used. Source data verification was 100%.

A multi-variate logistic regression analysis was conducted to assess known key risk factors for progression to IMV or death. Logistic regression models were built to predict Day 28 SWOV using known risk factors for progression to IMV or death that were available in the intent-to-treat (ITT) dataset. Three versions of the model were built: one with baseline ordinal score and not severity (stratification variable: severe or critical), one with severity and not baseline ordinal scale, and one with neither baseline ordinal scale nor severity. The set of covariates included in the models were:

- Treatment: lenzilumab or placebo
- Age ≥ 65 or <65 years
- Gender
- BMI: The value of BMI linear transformed to a scale where BMI 17=0.0, BMI 45=1.0
- Number of days before randomization of symptom onset (SYMDAY)
- Number of days before randomization of hospital admission (DIADAY)
- Baseline CRP
- Diabetes
- Heart condition: prior diagnosis of hypertension, coronary artery disease, or congestive heart failure
- Respiratory condition: prior diagnosis of asthma, COPD, or interstitial lung disease
- Vascular condition: prior diagnosis of cerebrovascular disorders or thrombosis and embolism
- Other risk factors: prior diagnosis of cancers (haematological or non-haematological), chronic kidney disease (including renal failure), chronic liver disease (including hepatic failure), or for being a smoker

Model type training was performed by bootstrapping, where 10,000 logistic regression models were built on random subsets of the ITT analysis set (n=520). For each bootstrapped model iteration, metrics were evaluated on the 20% test set and the feature coefficients of the model were recorded. This gave a distribution of 10,000 samples for the metrics and coefficients. All models produced similar outcomes. Therefore, the model chosen used severity as the covariate to be consistent with the covariate used in the pre-specified primary analysis, in addition to the other risk factors as covariates.

### Patient and public involvement

Patients were involved in this research. Members of the public were involved in the research only if they had a direct role in implementing the research or patient care. No other members of the public were involved in this work.

## RESULTS

### Demographics

Five hundred, twenty-eight participants were screened, of whom 520 were randomized and included in the ITT population (Figure 1).^13^ The mITT population represented 92% (479/520) of the total population, of which 90% and 94% of each population were randomized to lenzilumab (236/261) and placebo (243/259), respectively. Participants with CRP<150 mg/L comprised 73% of the mITT population (351/479) and 78.0% (351/450) of the mITT population with an evaluable baseline CRP. Baseline characteristics were well-balanced between treatment groups in CRP<150 and CRP>150 mg/L populations, as well as the overall mITT population (Table 1). No major differences were observed between these groups and these groups reflected the demographics of the overall population.

**Figure 1.**
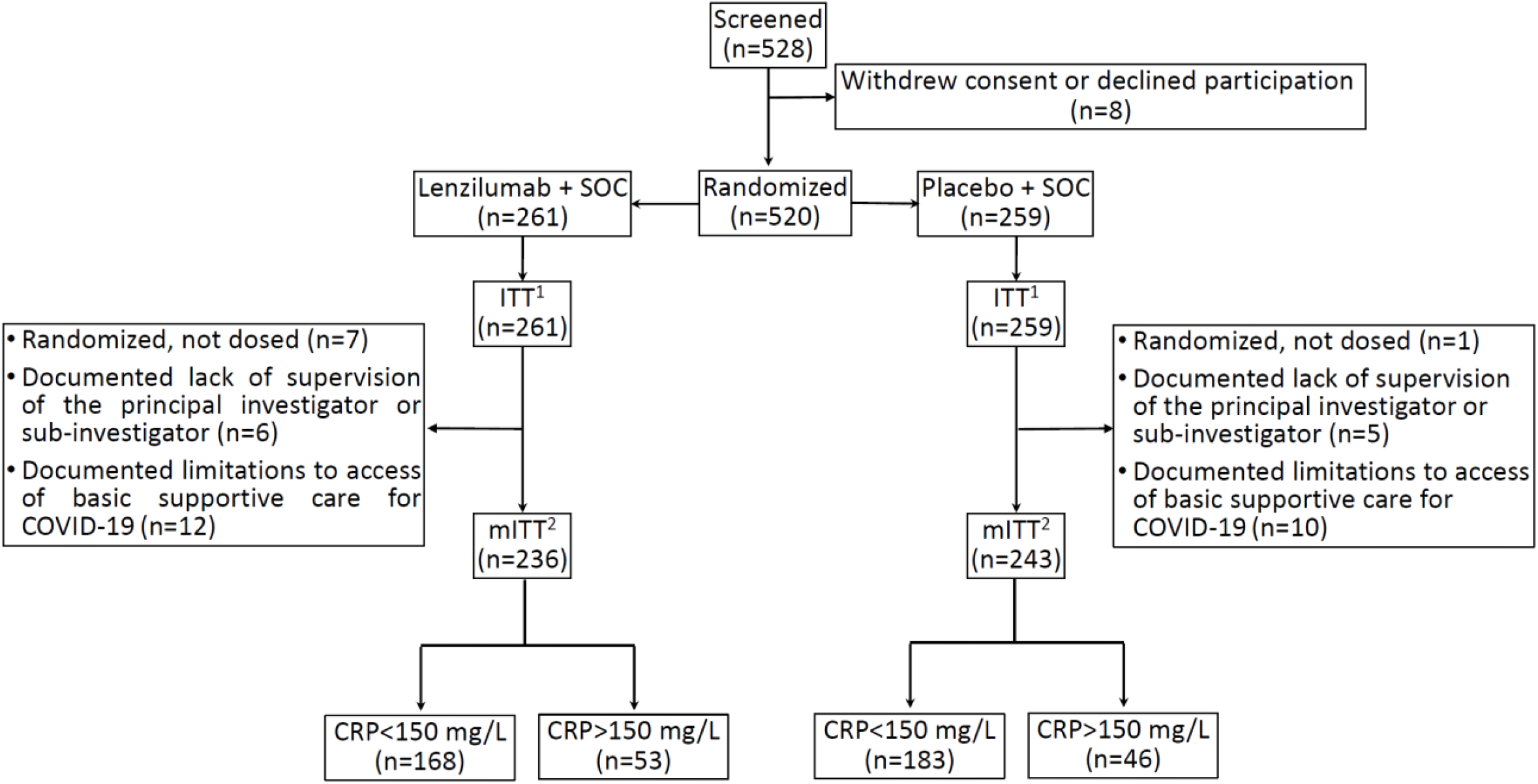
Randomization and Analysis Populations. The ITT population consisted of all randomized participants.^1^ The safety set included all participants who received at least one dose of study drug and is presented by the actual drug received.^2^ Randomized participants who received at least one dose of study drug under the documented supervision of the principal investigator or sub-investigator were included in the mITT population. This population excluded participants from sites that experienced documented limitations to access of basic supportive care for COVID-19.

**Table 1.**
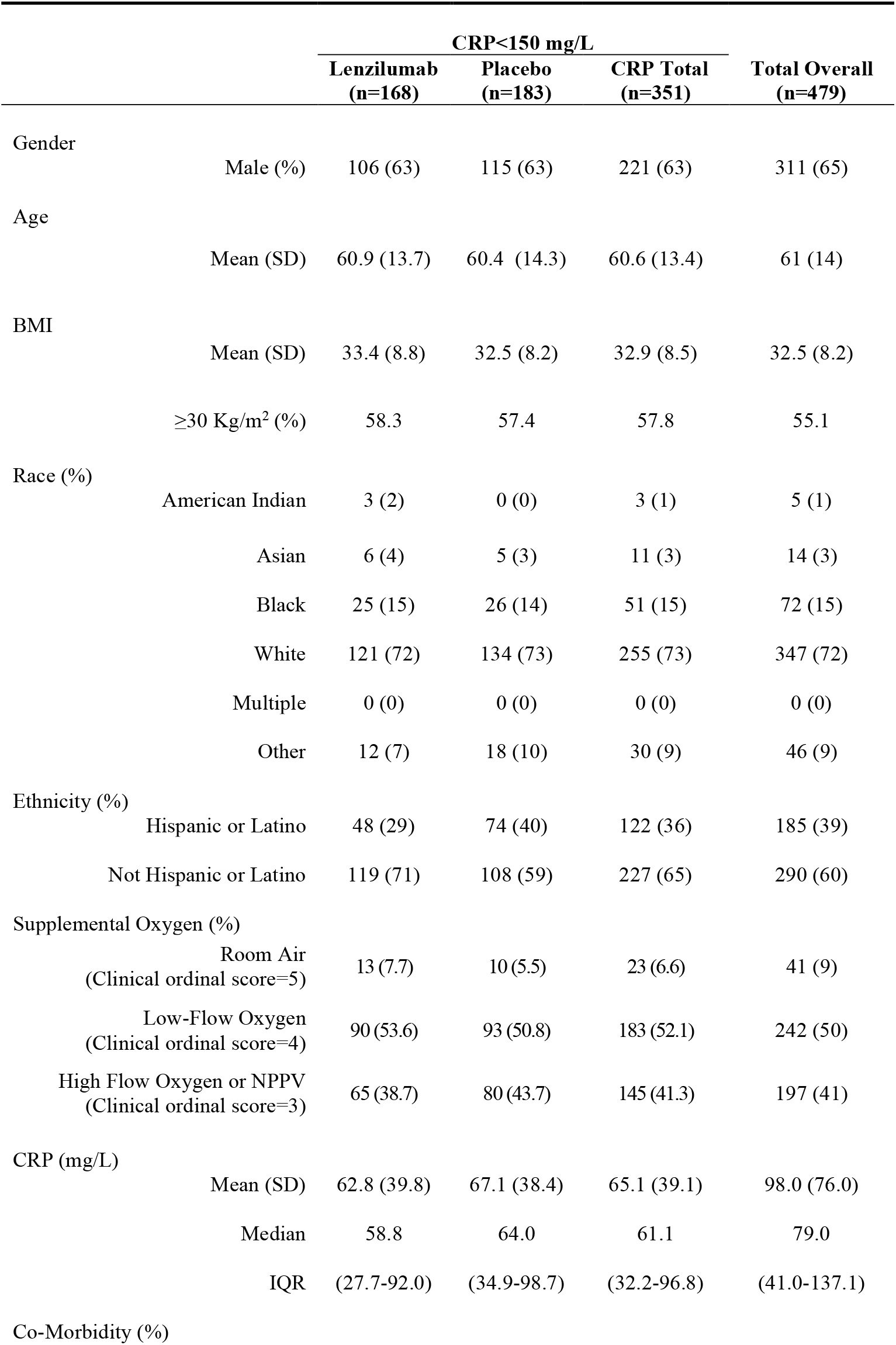
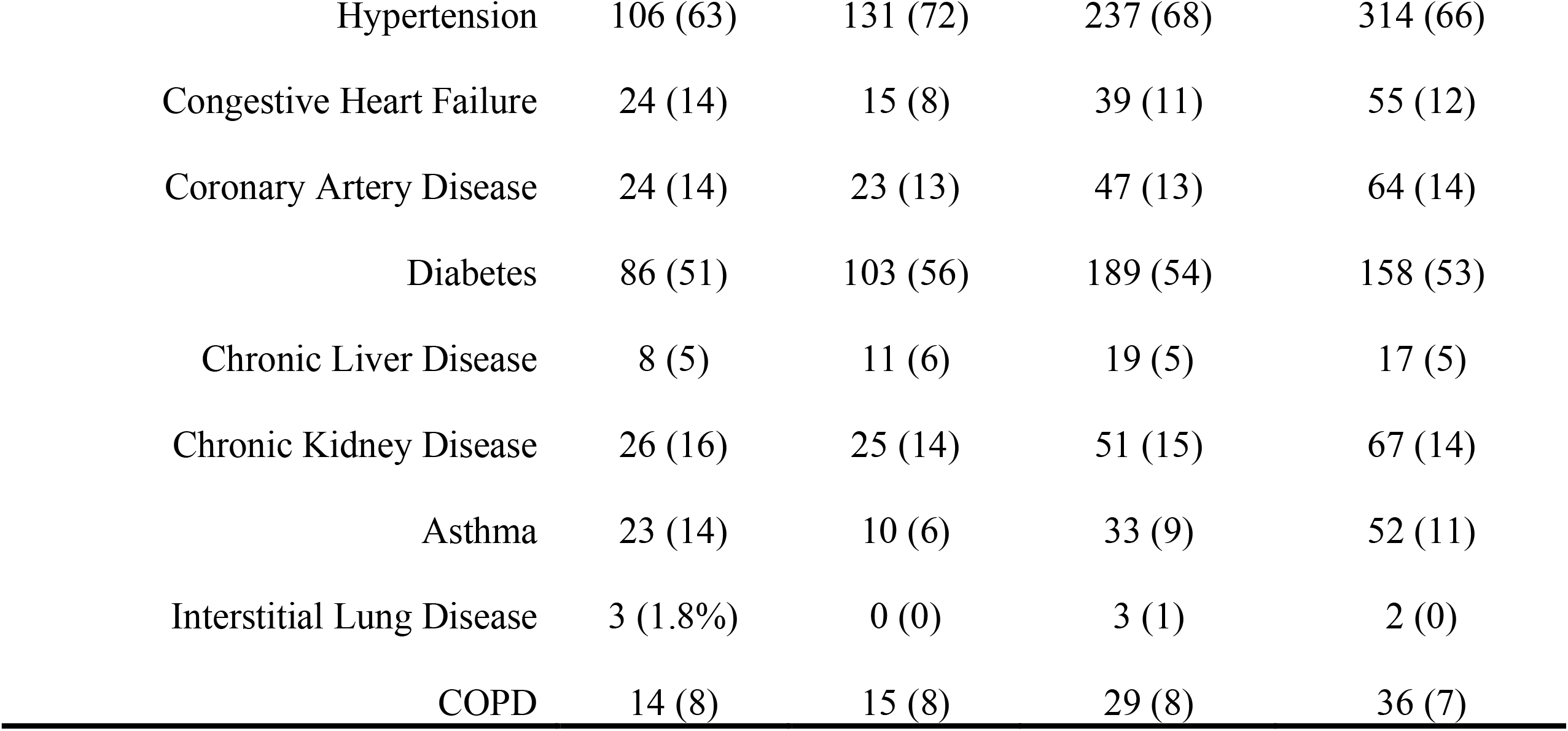
Baseline Characteristics.

|  |  | CRP<150 mg/L |  |  | Total Overall<br>(n=479) |
| --- | --- | --- | --- | --- | --- |
|  |  | Lenzilumab<br>(n=168) | Placebo<br>(n=183) | CRP Total<br>(n=351) |  |
| Gender |  |  |  |  |  |
|  | Male (%) | 106 (63) | 115 (63) | 221 (63) | 311 (65) |
| Age |  |  |  |  |  |
|  | Mean (SD) | 60.9 (13.7) | 60.4 (14.3) | 60.6 (13.4) | 61 (14) |
| BMI |  |  |  |  |  |
|  | Mean (SD) | 33.4 (8.8) | 32.5 (8.2) | 32.9 (8.5) | 32.5 (8.2) |
|  | ≥30 Kg/m<sup>2</sup> (%) | 58.3 | 57.4 | 57.8 | 55.1 |
| Race (%) |  |  |  |  |  |
|  | American Indian | 3 (2) | 0 (0) | 3 (1) | 5 (1) |
|  | Asian | 6 (4) | 5 (3) | 11 (3) | 14 (3) |
|  | Black | 25 (15) | 26 (14) | 51 (15) | 72 (15) |
|  | White | 121 (72) | 134 (73) | 255 (73) | 347 (72) |
|  | Multiple | 0 (0) | 0 (0) | 0 (0) | 0 (0) |
|  | Other | 12 (7) | 18 (10) | 30 (9) | 46 (9) |
| Ethnicity (%) |  |  |  |  |  |
|  | Hispanic or Latino | 48 (29) | 74 (40) | 122 (36) | 185 (39) |
|  | Not Hispanic or Latino | 119 (71) | 108 (59) | 227 (65) | 290 (60) |
| Supplemental Oxygen (%) |  |  |  |  |  |
|  | Room Air<br>(Clinical ordinal score=5) | 13 (7.7) | 10 (5.5) | 23 (6.6) | 41 (9) |
|  | Low-Flow Oxygen<br>(Clinical ordinal score=4) | 90 (53.6) | 93 (50.8) | 183 (52.1) | 242 (50) |
|  | High Flow Oxygen or NPPV<br>(Clinical ordinal score=3) | 65 (38.7) | 80 (43.7) | 145 (41.3) | 197 (41) |
| CRP (mg/L) |  |  |  |  |  |
|  | Mean (SD) | 62.8 (39.8) | 67.1 (38.4) | 65.1 (39.1) | 98.0 (76.0) |
|  | Median | 58.8 | 64.0 | 61.1 | 79.0 |
|  | IQR | (27.7-92.0) | (34.9-98.7) | (32.2-96.8) | (41.0-137.1) |
| Co-Morbidity (%) |  |  |  |  |  |

|  |  |  |  |  |
| --- | --- | --- | --- | --- |
| Hypertension | 106 (63) | 131 (72) | 237 (68) | 314 (66) |
| Congestive Heart Failure | 24 (14) | 15 (8) | 39 (11) | 55 (12) |
| Coronary Artery Disease | 24 (14) | 23 (13) | 47 (13) | 64 (14) |
| Diabetes | 86 (51) | 103 (56) | 189 (54) | 158 (53) |
| Chronic Liver Disease | 8 (5) | 11 (6) | 19 (5) | 17 (5) |
| Chronic Kidney Disease | 26 (16) | 25 (14) | 51 (15) | 67 (14) |
| Asthma | 23 (14) | 10 (6) | 33 (9) | 52 (11) |
| Interstitial Lung Disease | 3 (1.8%) | 0 (0) | 3 (1) | 2 (0) |
| COPD | 14 (8) | 15 (8) | 29 (8) | 36 (7) |

### Primary Outcome of LIVE-AIR

As reported previously, treatment with lenzilumab was associated with a greater likelihood of achieving SWOV compared to the placebo group (HR, 1·54; 95%CI, 1·02 to 2·32; p=0·0403; Table 2a, Figure 2a).^13^ The estimate of SWOV, through Day 28 was 198 (84%; 95%CI: 79 to 89) and 190 (78%; 72 to 83) in patients treated with lenzilumab or placebo, respectively. Separation of the survival curves occurred as early as 3 days following treatment (Figure 2a), continued to increase through approximately Day 10, and was maintained for the duration of the 28-day observation period. SWOV was also improved in those concomitantly administered remdesivir and corticosteroids^13^.

**Table 2a.** Primary Endpoint According to Baseline CRP^a,b^.

| Outcome | CRP < 150 mg/L (n=351) |  |  |  | Overall Population (N=479) |  |  |  | CRP ≥ 150 mg/L (n=99) |  |  |  |
| --- | --- | --- | --- | --- | --- | --- | --- | --- | --- | --- | --- | --- |
|  | Median CRP, 61 mg/L; IQR (32 to 97 mg/L) |  |  |  | Median CRP, 79 mg/L; IQR (41 to 137 mg/L) |  |  |  | Median CRP, 201 mg/L; IQR (175 to 32 mg/L) |  |  |  |
|  | Lenzilumab<br>(n=168) | Placebo<br>(n=183) | HR or OR<br>(95%CI) | p value | Lenzilumab<br>(n=236) | Placebo<br>(n=243) | HR or OR<br>(95%CI) | p value | Lenzilumab<br>(n=53) | Placebo<br>(n=46) | HR or OR<br>(95%CI) | p value |
| **SWOV**<br>**(%; 95%CI)** | 152 (90)<sup>e</sup><br>(85 to 94) | 144 (79)<sup>e</sup><br>(72.1 to 84.1) | 2.54<sup>f</sup><br>(1.46 to 4.41) | 0.0009 | 198 (84)<sup>e</sup><br>(79 to 89) | 190 (78)<sup>e</sup><br>(72 to 83) | 1.54<sup>f</sup><br>(1.02-2.32) | 0.0403 | 37 (69.3)<sup>e</sup><br>(55 to 80) | 33 (72)<sup>e</sup><br>(56 to 83) | 1.04<sup>f</sup><br>(0.51 to 2.14) | 0.9058 |

**Table 2b.** Key Secondary Endpoints According to Baseline CRP^a,b^.

| Outcome | CRP < 150 mg/L (n=351) |  |  |  | Overall Population (N=479) |  |  |  | CRP ≥ 150 mg/L (n=99) |  |  |  |
| --- | --- | --- | --- | --- | --- | --- | --- | --- | --- | --- | --- | --- |
|  | Median CRP, 61 mg/L; IQR (32 to 97 mg/L) |  |  |  | Median CRP, 79 mg/L; IQR (41 to 137 mg/L) |  |  |  | Median CRP, 201 mg/L; IQR (175 to 232 mg/L) |  |  |  |
|  | Lenzilumab<br>(n=168) | Placebo<br>(n=183) | HR or OR<br>(95%CI) | p value | Lenzilumab<br>(n=236) | Placebo<br>(n=243) | HR or OR<br>(95%CI) | p value | Lenzilumab<br>(n=53) | Placebo<br>(n=46) | HR or OR<br>(95%CI) | p value |
| **IMV, ECMO, or**<br>**Mortality**<br>**% (95%CI)** | 14 (8)<sup>c</sup><br>(5 to 14) | 34 (19)<sup>c</sup><br>(13 to 26) | 0.38<sup>d</sup><br>(0.19 to 0.75) | 0.0053 | 35 (15)<sup>c</sup><br>(11 to 21) | 51 (21)<sup>c</sup><br>(16 to 27) | 0.67<sup>d</sup><br>(0.41-1.10) | 0.111 | 19 (30)<sup>c</sup><br>(19 to 44) | 12 (27)<sup>c</sup><br>(16 to 43) | 1.14<sup>d</sup><br>(0.46 to 2.86) | 0.7731 |
| **IMV**<br>**% (95%CI)** | 10 (6)<sup>e</sup><br>(3 to 11) | 36 (20)<sup>e</sup><br>(14 to 26) | 0.28<sup>f</sup><br>(0.15 to 0.54) | 0.0002 | 11.0<sup>e</sup><br>(8-16) | 42.0<sup>e</sup><br>(16-26) | 0.52<sup>f</sup><br>(0.32-0.82) | 0.0059 | 13 (24)<sup>e</sup><br>(14 to 38) | 13 (28)<sup>e</sup><br>(17 to 44) | 0.77<sup>f</sup><br>(0.34 to 1.68) | 0.5098 |
| **Mortality**<br>**% (95%CI)** | 13 (8)<sup>e</sup><br>(5 to 13) | 26 (14)<sup>e</sup><br>(10 to 20) | 0.57<sup>f</sup><br>(0.29-1.12) | 0.1040 | 24 (10)<sup>e</sup><br>(6 to 14) | 34 (14)<sup>e</sup><br>(10 to 19) | 0.72<sup>f</sup><br>(0.42-1.23) | 0.2410 | 7 (13)<sup>e</sup><br>(7 to 26) | 6 (13)<sup>e</sup><br>(6 to 27) | 0.88<sup>f</sup><br>(0.29 to 2.63) | 0.8165 |
| **Ventilator-Free**<br>**Days mean (SD)** | 25.7<br>(7.6) | 22.7<br>(10.5) |  | 0.0045<sup>g</sup> | 24.5<br>(8.8) | 22.6<br>(10.5) |  | 0.0766<sup>g</sup> | 20.8<br>(11.2) | 21.7<br>(10.6) |  | 0.83<sup>g</sup> |
| **ICU Days**<br>**Mean (SD)** | 3.9<br>(8.3)<br>(n=168) | 6.2<br>(10.6)<br>(n=183) |  | 0.0458<sup>g</sup> | 5.4<br>(9.6) | 6.6<br>(10.7) |  | 0.1604<sup>g</sup> | 9.6<br>(11.5) | 8.5<br>(11.2) |  | 0.9400<sup>g</sup> |
| **Time to Recovery**<br>**(days)** |  |  |  |  |  |  |  |  |  |  |  |  |
| **Quartile** |  |  |  |  |  |  |  |  |  |  |  |  |
| **25%** | 4<br>(4,5) | 5<br>(5,5) |  |  | 5<br>(4,5) | 8<br>(5,5) |  |  | 8<br>(6-9) | 6<br>(5-8) |  |  |
| **50%** | 7<br>(6-8) | 8<br>(7-9) |  | 0.0219<sup>f</sup> | 8<br>(7-9) | 8<br>(7-9) |  | 0.432<sup>f</sup> | 12<br>(9-19) | 11<br>(7-18) |  | 0.153<sup>f</sup> |
| **75%** | 11<br>(10-14) | 17<br>(12-NA) |  |  | 15<br>(11-20) | 19<br>(13-NA) |  |  | NA<br>(19-NA) | NA<br>(17-NA) |  |  |

<sup>a</sup> All data censored at 28 days following enrollment.
<sup>b</sup> mITT, modified intention to treat population
<sup>c</sup> Estimated marginal mean
<sup>d</sup> Odds Ratio with age (≤65, >65) and severity (severe, critical) strata as covariates
<sup>e</sup> Kaplan-Meier estimates for proportion of participants
<sup>f</sup> Cox Proportional Hazard Model for time to event with age (≤65, >65) and severity (severe, critical) strata as covariates
<sup>g</sup> Stratified Wilcoxon Rank Sum Test with age (≤65, >65) and severity (severe, critical) strata as covariates

**Figure 2.**
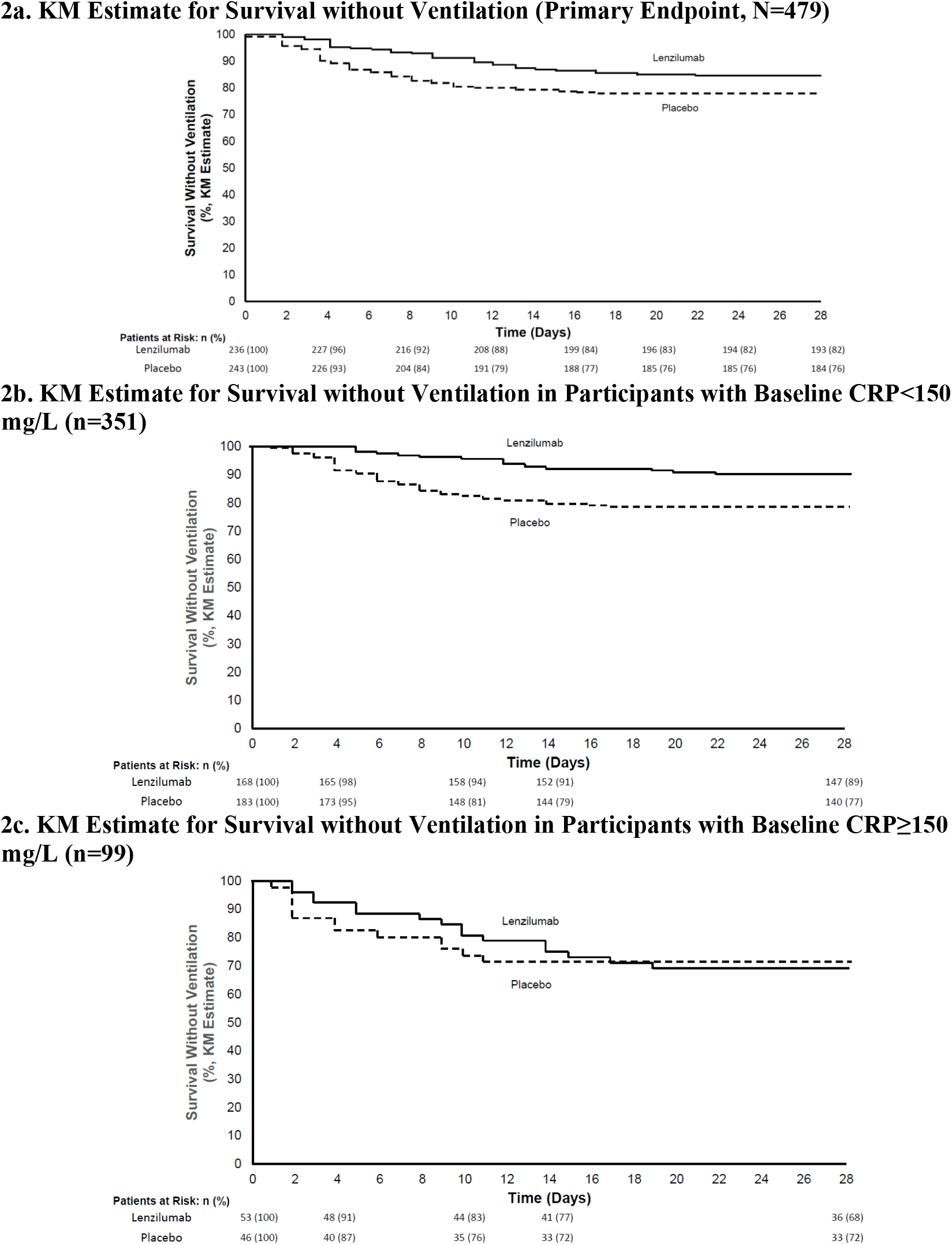
Kaplan-Meier Estimate for Survival Without Ventilation. 2**a**. KM Estimate for Survival Without Ventilation (Primary Endpoint). The primary efficacy analysis was based on the overall mITT population. Separation of the survival curves occurred as early as 3 days following treatment. Following Day 10, separation maintained for the duration of the observation period. Lenzilumab treatment improved the relative likelihood of achieving SWOV compared with placebo (HR: 1.54; 95%CI: 1.02-2.32, p=0.0403). Reprinted from Lancet Respiratory Medicine. Temesgen Z, Burger CD, Baker J, Polk C, Libertin CR, Kelley CF, Marconi VC, Orenstein R, Catterson VM, Aronstein WS, Durrant CD, Chappell, D, Ahmed O, Chappell G, Badley AD, for the LIVE-AIR Study Group, Lenzilumab in hospitalised patients with COVID-19 pneumonia (LIVE-AIR): a phase 3, randomised, placebo-controlled trial, DOI:https://doi.org/10.1016/S2213-2600(21)00494-X, Copyright (2021), with permission from Elsevier. **2b**. KM Estimate for Survival Without Ventilation in Participants with baseline CRP<150 mg/L. Separation of the survival curves occurred after two days post treatment. The separation of the curves were more pronounced than in the overall mITT analysis. Lenzilumab treatment improved the relative likelihood of achieving SWOV compared with placebo (HR: 2.54; 95%CI: 1.46-4.41, p=0.0009). **2c**. KM Estimate for Survival Without Ventilation in Participants with baseline CRP≥150 mg/L.

### Risk Factors Affecting SWOV in LIVE-AIR

Twelve risk factors were evaluated for their influence on SWOV. Incorporating these known risk factors as covariates into an iterative multivariate logistic regression analysis demonstrated a statistically significant positive outcome for SWOV with lenzilumab treatment (OR, 1.51; 95%CI, 1.18 to 1.94; nominal p=0.0006; Figure 3). This model also demonstrated that elevated baseline plasma CRP was the most predictive factor for progression to IMV or death (OR, 0.15; 95%CI, 0.07 to 0.29; nominal p<0.001; Figure 3).

**Figure 3.**
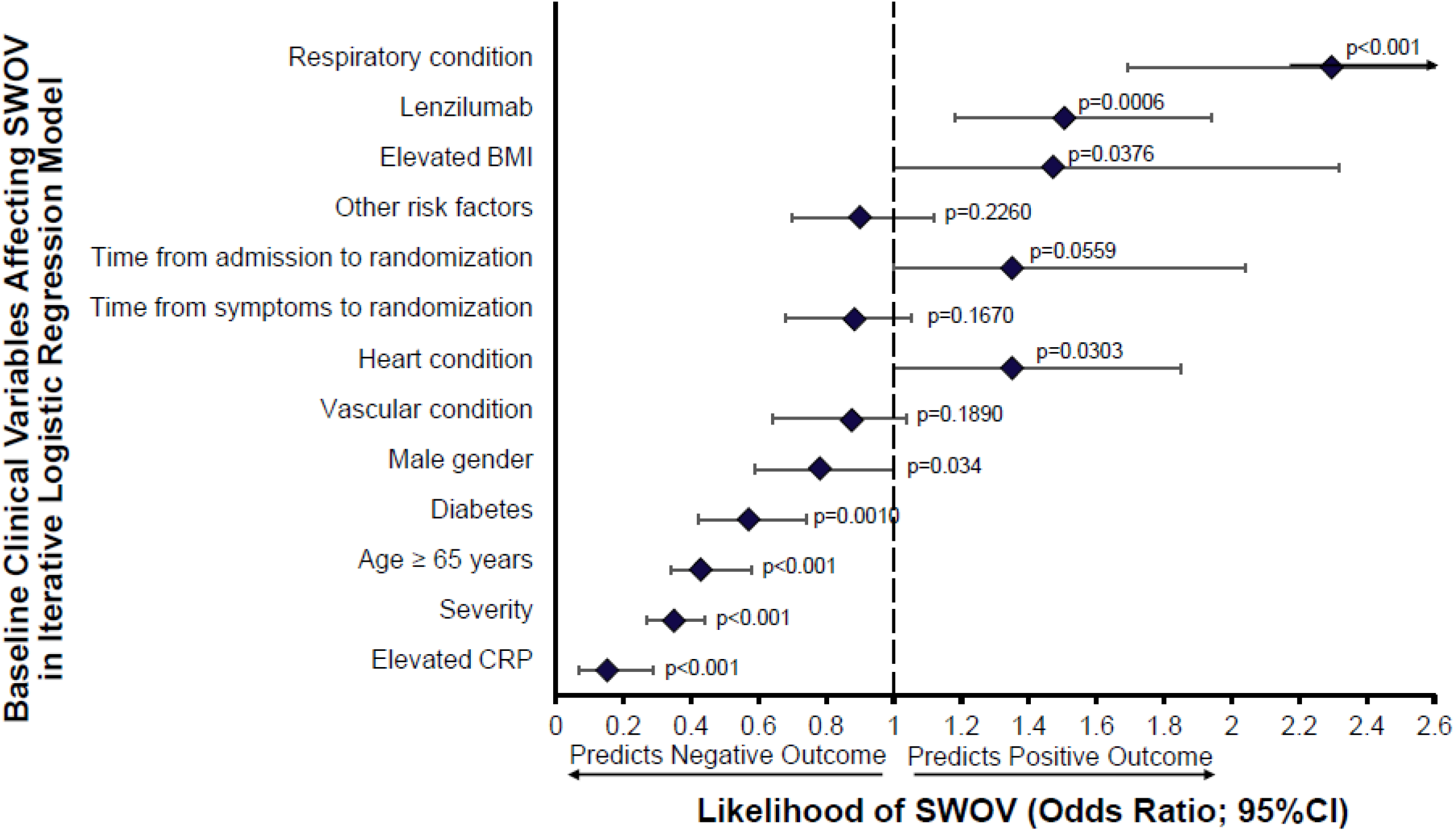
Impact of Baseline Demographics and Risk Factors on Survival with Ventilation Using an Iterative Multivariate Logistic Regression Model. Twelve covariates were included in the model encompassing known risk factors for progression to IMV and/or death by Day 28: Baseline CRP (CRP), disease severity at randomization (severity), respiratory condition (asthma, COPD, interstitial lung disease), age >=65, diabetes (Type 1 or Type 2), lenzilumab (treated or placebo), BMI, time from admission to randomization, time from symptoms to randomization, heart condition (hypertension, coronary artery disease, congestive heart failure), male gender, vascular condition (cerebrovascular disorders, thrombosis or embolism), other risk factors (prior diagnosis of cancer; haematological or non-haematological), chronic kidney disease (including renal failure), or chronic liver disease (including hepatic failure). Statistical significance was reached for all features with a displayed p-value.

LIVE-AIR was not stratified by baseline CRP level nor was CRP a covariate in any of the pre-specified outcome measures. The *post hoc* inclusion of CRP as a covariate in the overall mITT analysis population, along with age and disease severity, resulted in a statistically significant lenzilumab treatment effect on SWOV (HR: 1.74; 95%CI, 1.14 to 2.66; p=0.0101) as well as several key secondary endpoints, including incidence of IMV, ECMO, or death (OR: 0.55; 95%CI, 0.32 to 0.94; p=0.029) and ventilator-free days (mean 24.5 vs. 22.6, p=0.021). Further analysis demonstrated a significant statistical interaction between lenzilumab treatment and CRP (p=0.044).

An exploratory analysis for the effect of lenzilumab on SWOV was conducted by CRP baseline quartile. Response to lenzilumab was observed in the first through third quartiles of baseline CRP with the greatest lenzilumab treatment effect observed in the first quartile (CRP<41 mg/L; HR: 8.20; 95%CI; 1.74 to 38.69; p=0.0079) and a numeric difference that did not reach statistical significance in the second quartile and a significant treatment effect observed in the third quartile (CRP 79<137 mg/L; HR: 2.25; 95%CI; 1.04 to 4.88; p=0.0407 (Table 3).

**Table 3.**
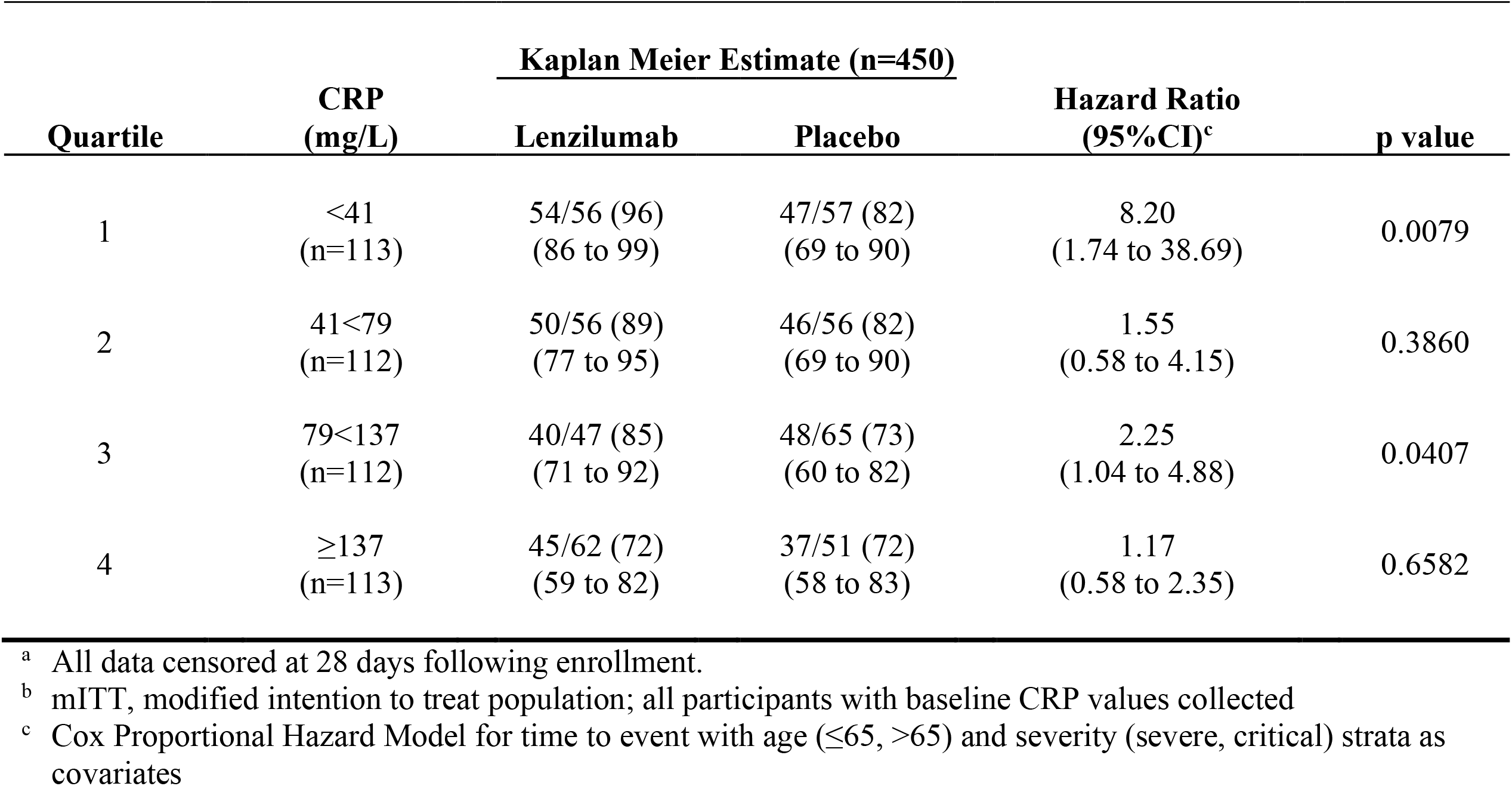
Analysis of Treatment on SWOV According to Baseline CRP Quartile.^a,b^.

| Quartile | CRP<br>(mg/L) | Kaplan Meier Estimate (n=450) |  | Hazard Ratio<br>(95%CI)<sup>c</sup> | p value |
| --- | --- | --- | --- | --- | --- |
|  |  | Lenzilumab | Placebo |  |  |
| 1 | <41<br>(n=113) | 54/56 (96)<br>(86 to 99) | 47/57 (82)<br>(69 to 90) | 8.20<br>(1.74 to 38.69) | 0.0079 |
| 2 | 41<79<br>(n=112) | 50/56 (89)<br>(77 to 95) | 46/56 (82)<br>(69 to 90) | 1.55<br>(0.58 to 4.15) | 0.3860 |
| 3 | 79<137<br>(n=112) | 40/47 (85)<br>(71 to 92) | 48/65 (73)<br>(60 to 82) | 2.25<br>(1.04 to 4.88) | 0.0407 |
| 4 | ≥137<br>(n=113) | 45/62 (72)<br>(59 to 82) | 37/51 (72)<br>(58 to 83) | 1.17<br>(0.58 to 2.35) | 0.6582 |

<sup>a</sup> All data censored at 28 days following enrollment.
<sup>b</sup> mITT, modified intention to treat population; all participants with baseline CRP values collected
<sup>c</sup> Cox Proportional Hazard Model for time to event with age (≤65, >65) and severity (severe, critical) strata as covariates

Given the greatest treatment effect for lenzilumab was observed in the first through third quartiles, an analysis of baseline plasma CRP levels and the likelihood to achieve SWOV with lenzilumab was further explored at baseline CRP greater than 100 mg/L (Figure 4). This CRP level and the 25 mg/L increments explored were arbitrarily selected with the knowledge that the highest quartile value for baseline CRP levels was ≥ 137 mg/L. In this analysis, the hazard ratio for SWOV was calculated for all cumulative participants with CRP levels below the indicated cutoff value. The lenzilumab treatment effect and baseline CRP level demonstrated a sigmoidal relationship. The hazard ratio resulting from lenzilumab treatment was above 2.25 for baseline CRP levels between 100 and 150 mg/L, and progressively declined above 150 mg/L until 275 mg/L where the HR plateaued at approximately 1.5.

**Figure 4.**
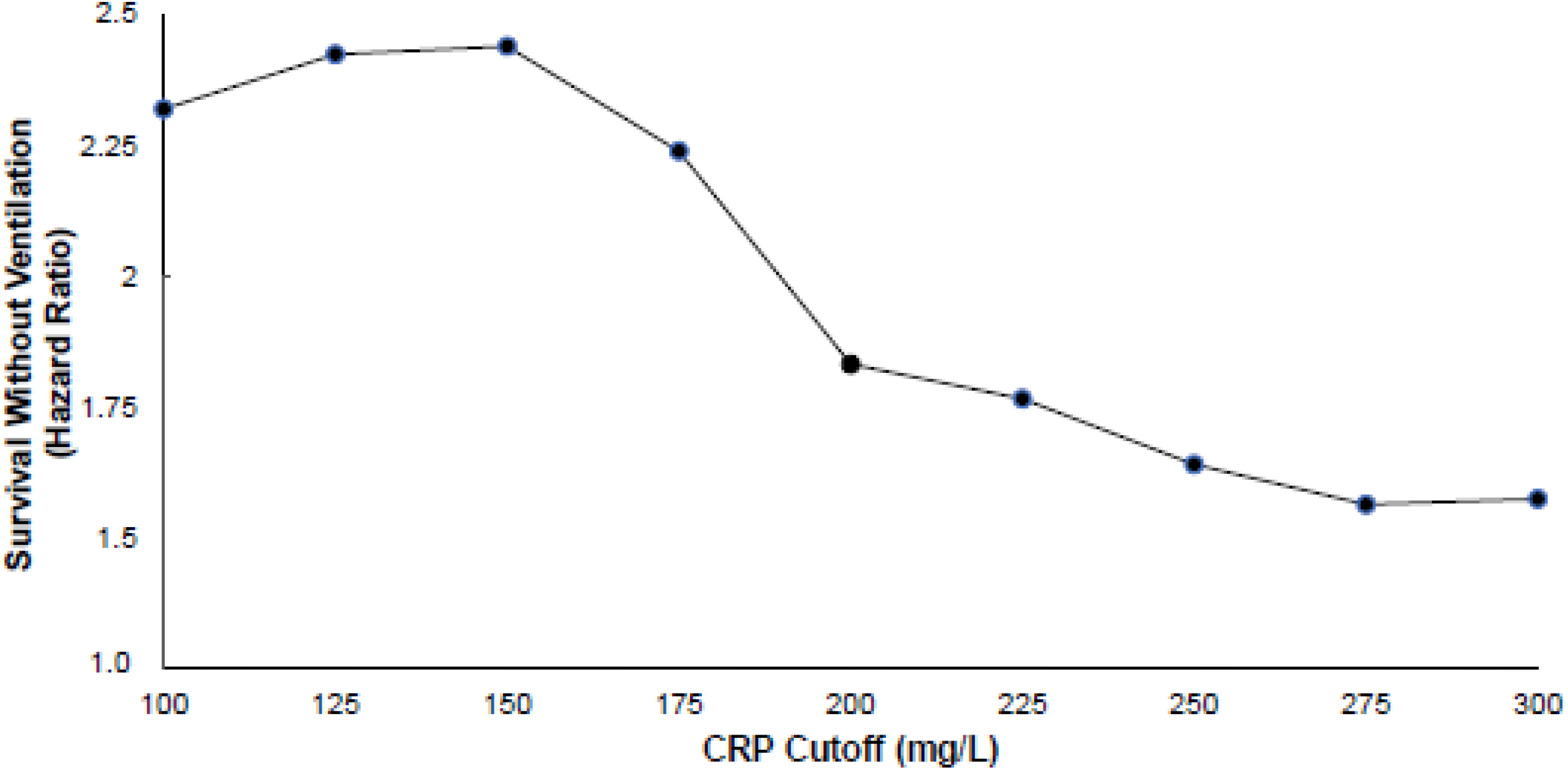
Likelihood of Survival Without Ventilation by Level of CRP Cutoff. The hazard ratio for SWOV was calculated for all participants, with CRP level below the indicated cutoff value. Participants with CRP<150 mg/L had the greatest likelihood of achieving SWOV.

### Effect of CRP<150 mg/L on SWOV and Secondary Endpoints in LIVE-AIR

In participants with baseline CRP<150 mg/L, lenzilumab improved the likelihood of SWOV compared with placebo (HR: 2.54; 95%CI; 1.46 to 4.41; nominal p=0.0009; Table 2b, Figure 2b). Separation of the survival curves appeared earlier than in the overall population and followed a similar pattern as the overall population thereafter (Figure 2b). SWOV, in response to lenzilumab treatment, was similar to placebo in participants with CRP≥150 mg/L at baseline. (Table 2 and Figure 2c).

### Secondary Outcomes

Secondary outcomes were improved with lenzilumab treatment in participants with CRP<150 mg/L. Incidence of IMV, ECMO or death with lenzilumab treatment was not statistically improved in the overall mITT population but was less in participants with baseline CRP<150 mg/L (OR 0.38; 0.19 to 0.75; nominal p=0.0053; Table 2b). Additional secondary endpoints were improved with lenzilumab treatment in participants with baseline CRP<150 mg/L (Table 2b). Ventilator-free days were 25.7 (SD: 7.6) and 22.7 (10.5) with lenzilumab or placebo treatment, respectively (nominal p=0.0045). This difference was not observed with baseline CRP≥150 mg/L. ICU days were also less with lenzilumab compared with placebo treatment in participants with baseline CRP<150 mg/L (nominal p=0.0458). Time to recovery with lenzilumab treatment was improved with lenzilumab treatment relative to placebo in participants with a baseline CRP<150mg/L (p=0.0219).

The LIVE-AIR trial was not powered to demonstrate a mortality benefit. The likelihood of mortality was numerically lowest in baseline CRP<150 mg/L but did not reach statistical significance (HR:0.57; 95%CI, 0.29 to 1.12; p=0.104).

### Time Course of Changes in CRP

In the overall mITT population regardless of treatment assignment, baseline CRP levels were related to COVID-19 severity at baseline. CRP levels at baseline increased with ordinal scale where participants on room air exhibited average CRP levels of 83.6 (SE: 11.8) mg/L; low flow O_2_, 95.2 (4.5); and high flow O_2_, 104.0 (5.9).

In participants who required IMV or died, mean CRP levels were elevated and remained so through Day 28 compared to participants who achieved SWOV (Figure 5a). The mean CRP time course in participants who achieved SWOV rapidly decreased from baseline through Day 4 and remained low through Day 28. The CRP level at baseline for participants who required IMV or died was 128.5 (SE: 86.2) mg/L compared to 91.2 (71.1) mg/L in those who achieved SWOV. For those participants who required IMV or died, CRP level within ±1 day of the event was 178 (52.4) mg/L (median: 167 mg/L). Mean CRP>100 mg/L during the hospital course was associated with all events of IMV and/or death in the trial, whereas mean CRP was <50 mg/L during the hospital course in participants who achieved SWOV. Participants in the placebo arm with baseline CRP>150 mg/L progressed to IMV and death with time to event in the 25^th^ and 50^th^ percentiles of 2 and 4 days respectively. Those with CRP>150 mg/L at any time are at significant risk of an event, accounting for 72% of all failures to achieve SWOV in LIVE-AIR.

**Figure 5.**
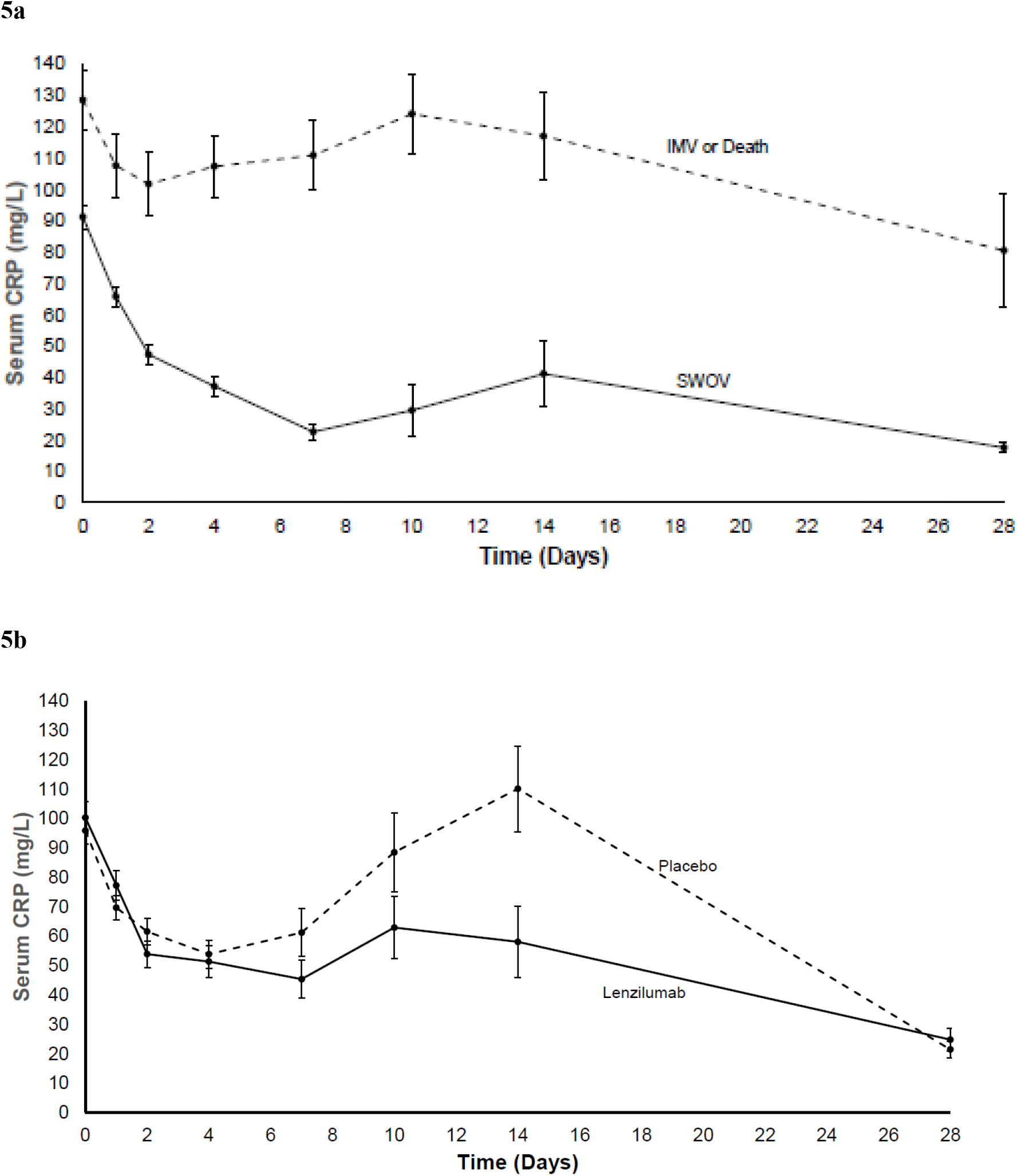
Analysis of CRP Levels Over Time through Day 28. **5a. CRP Levels Over Time in Participants who met Primary Endpoint (SWOV) vs. Participants who Progressed to IMV and/or Death**. This analysis was conducted on the entire mITT population without regard to treatment. CRP levels in participants requiring IMV or who died remained elevated through the hospital course. CRP level were lower in participants who achieved SWOV. **5b. CRP Levels Over Time in Participants Treated with Lenzilumab vs. Placebo**. Lenzilumab decreased plasma CRP levels relative to placebo by day 7 and through day 14 following treatment. (Values are mean ± standard error; mITT population).

CRP levels were reduced by lenzilumab treatment (Figure 5b). By Day 2 following lenzilumab treatment, mean CRP levels were lower in than in the placebo group. CRP levels remained lower throughout the study until day of discharge or Day 28 when mean CRP recovered regardless of treatment.

### Safety

In the safety population, adverse events ≥ Grade 3 were reported in 18% of the participants treated with lenzilumab and 28% of participants treated with placebo in those with baseline CRP<150 mg/L (Table 4). Respiratory, thoracic, and mediastinal disorders were less common in the lenzilumab group with CRP<150 mg/L relative to placebo. The differences in this group were driven mostly by a lower incidence of respiratory failure and acute respiratory failure associated with lenzilumab treatment. Additionally, infections and infestations, vascular disorders and renal and urinary disorders, and general and administration site disorders were all lower in the lenzilumab group with CRP <150 mg/L relative to placebo. No infusion-related reactions or serious adverse events; including, haematologic laboratory abnormalities, liver enzyme abnormalities, increased incidence of infection, or cases of pulmonary alveolar proteinosis were reported with lenzilumab treatment.

**Table 4.** Most Common Grade ≥3 Adverse Events (Overall Incidence ≥ 1.0%)

| System Organ Class<br>Preferred term n (%) | CRP<150 mg/L |  |  | Total Overall<br>(n=512) |
| --- | --- | --- | --- | --- |
|  | Lenzilumab<br>(n=181) | Placebo<br>(n=193) | Total<br>(n=374) |  |
| Any AE ≥ Grade 3 | 32 (17) | 54 (28) | 86 (23) | 152 (30) |
| Respiratory, thoracic, and mediastinal disorders | 30 (17) | 48 (25) | 78 (21) | 135 (26) |
| Respiratory Failure | 9 (5) | 22 (11) | 31 (8) | 54 (11) |
| Acute respiratory failure | 8 (4) | 15 (8) | 23 (6) | 40 (8) |
| Hypoxia | 9 (5) | 10 (5) | 19 (5) | 30 (6) |
| Pulmonary embolism | 2 (1) | 3 (2) | 5 (1) | 8 (2) |
| Cardiac disorders | 7 (4) | 10 (5) | 17 (5) | 29 (6) |
| Cardiac arrest | 5 (3) | 3 (2) | 8 (2) | 12 (2) |
| Acute Myocardial Infarction | 0 (0) | 3 (2) | 3 (1) | 3 (1) |
| Vascular disorders | 4 (2) | 8 (4) | 12 (3) | 25 (5) |
| Shock | 1 (1) | 3 (2) | 4 (1) | 9 (2) |
| Infections and infestations | 4 (2) | 7 (4) | 11 (3) | 26 (5) |
| Septic Shock | 4 (2) | 5 (3) | 9 (2) | 14 (3) |
| Sepsis | 0 (0) | 3 (2) | 3 (1) | 7 (1) |
| General disorders and administration site conditions | 2 (1) | 8 (4) | 10 (3) | 12 (2) |
| Multiple organ dysfunction syndrome | 2 (1) | 6 (3) | 8 (2) | 9 (2) |
| Renal and urinary disorders | 1 (1) | 7 (4) | 8 (2) | 16 (3) |
| Acute kidney injury | 1 (1) | 4 (2) | 5 (1) | 13 (3) |
| Nervous system disorders | 0 (0) | 3 (2) | 3 (1) | 4 (1) |
| General disorders | 0 (0) | 3 (2) | 3 (1) | 3 (1) |

## DISCUSSION

Lenzilumab significantly improved SWOV in adults hospitalized with COVID-19 pneumonia compared to placebo. This improvement was most marked in participants with baseline CRP<150 mg/L. Incidence of IMV, ECMO or death, ventilator free days, ICU days, and time to recovery were also significantly improved in participants with a baseline CRP<150 mg/L who received lenzilumab compared to placebo. When baseline risk factors were analyzed in a multivariate model for their impact on SWOV, lenzilumab was a significant predictor of SWOV and baseline CRP was the greatest predictor of IMV and death. Response to lenzilumab was observed in the first through third quartiles of baseline CRP. Patients that progressed to IMV or death had elevated mean CRP levels through the hospital course. While baseline CRP levels were associated with COVID-19 severity at baseline and the likelihood of achieving SWOV regardless of treatment allocation, lenzilumab decreased CRP more rapidly than placebo and to levels more predictive of SWOV. Lenzilumab was well tolerated with no attributable serious adverse events.

Utilization of CRP as a biomarker of the extent of hyperinflammatory immune response to guide treatment in COVID-19 is supported by numerous reports and aligns with the immuno-pathophysiology as described herein. Elevation of CRP is driven by IL-6, ^5,6^ a downstream pro-inflammatory effector cytokine of hyperinflammatory immune response^16^ resulting from GM-CSF production. GM-CSF itself is elevated early in the hyperinflammatory immune response of COVID-19 and is associated with increased severity and poor outcomes.^1,17^ In LIVE-AIR, participants that progress to IMV or death had mean CRP values consistently above 100 mg/L during their hospital course. Seventy-two percent of participants who progressed to IMV and/or death in LIVE-AIR had CRP>150 mg/L at some point during their hospitalization and those with CRP>150 mg/L at baseline required rapid escalation of respiratory care within 2 to 4 days. The LIVE-AIR results confirm previous reports that elevated CRP (>150 mg/L) is predictive of the imminent risk of IMV or death.^1,10^ Taken together, the evidence suggests that lenzilumab interferes with GM-CSF signaling resulting in prevention of the multiplicity of downstream cytokine release, including IL-6, which leads to elevated CRP levels.^5,6,18^ This also explains why improvements in both primary and secondary endpoints were not seen in participants that had baseline CRP>150 mg/L. This level of CRP may reflect stages of hyperinflammatory immune response in which sufficient myeloid activation was already ongoing for GM-CSF neutralization to adequately prevent disease progression.

Recent published evaluations have begun to suggest COVID-19 patient phenotypes that may benefit most from various treatments. The IL-6 receptor blocker tocilizumab improved outcomes in patients with more advanced COVID-19 disease with median baseline CRP of 143 mg/L.^19,20^ Separately, tocilizumab decreased the risk of death and ICU admission or death among patients with baseline CRP>150 mg/L but not among those with baseline CRP≤ 150 mg/L.^21,22^ Tocilizumab is now recommended for use in ICU patients who require IMV or have rapidly increasing oxygen demands and have CRP >75 mg/L.^23^ While the temporal relationship between pro-inflammatory cytokines and CRP is likely complex, the use of CRP levels to guide treatment selection is emerging. In patients with CRP≥200mg/L, systemic glucocorticoids, administered within 48 hours of admission, were most effective in reducing progression to IMV and/or death compared to control (adjusted OR: 0.20; 95%CI: 0.05-0.67); however, in patients with CRP<99 mg/L systemic corticosteroid use caused harm (adjusted OR: 3.14; 95%CI: 1.52-6.50).^11^ Other clinical makers have also been associated with positive treatment effects. A four-phase model of progressive COVID-19 severity has been postulated from clinical experience based on objective endpoints (including CRP), combined with preclinical rationale, to propose use of anti-spike monoclonal and anti-GM-CSF antibodies in less severe COVID-19 and direct dexamethasone, anti-IL-6 antibodies, and JAK inhibitors for use in more advanced disease. ^12^

Therefore, inhibition of GM-CSF signaling, guided using CRP as a biomarker for emerging hyperinflammatory immune response, and prior to excessive elevations in CRP (i.e., >150 mg/L), may be an opportune therapeutic approach to prevent progression to advanced disease. GM-CSF activity could hypothetically fit into the recently proposed four phase model. ^12^ Elevation in GM-CSF may occur during the “early treatment phase”, referred to as phase 2, when viral replication and symptoms of emerging hyperinflammatory immune response are evident. The pro-inflammatory cytokine cascade during this phase is consistent with GM-CSF orchestrated myeloid activation and may be when GM-CSF neutralization is most effective. Accordingly, JAK inhibitors, corticosteroids, and anti-IL-6 monoclonal antibodies are proposed in the “dyspnea to ARDS” phase (phase 3) and the “ARDS” phase (Phase 4) where their activity on targets downstream from GM-CSF may have greater utility. ^12^

Limitations are associated with the analytic approach herein. The exploratory analysis of CRP as it relates to the primary endpoint of the likelihood of achieving SWOV was pre-specified, all other analyses were *post-hoc*, and none were prospectively powered. Therefore, the results should be interpreted with this caveat in mind. The findings herein will be further evaluated in the NIH-sponsored ACTIV-5/BET-B trial, that includes lenzilumab and where the primary efficacy analysis prospectively evaluates incidence of IMV, ECMO, or death in participants with baseline CRP<150 mg/L.

In summary, this comprehensive analysis of LIVE-AIR CRP data provides evidence for the utility of CRP to predict progression to IMV and death. GM-CSF neutralization with lenzilumab significantly improved SWOV in adults hospitalized with COVID-19 pneumonia compared to placebo. Those participants who had baseline CRP levels <150 mg/L responded more favorably to lenzilumab treatment, than those with CRP>150 mg/L. These finding suggest that CRP may be a useful biomarker in determining which participants may be most successfully treated with lenzilumab.

## Data Availability

All data produced in the present work are contained in the manuscript

## ACKNOWLEDGMENTS

The LIVE-AIR study team thanks and acknowledges all participants for their participation in this important research; CTI Clinical Trial & Consulting Services for their support as the clinical research organization managing the operations of the study; RxMedical Dynamics, LLC; and BioSymetrics, Inc.

## CONFLICT OF INTEREST

ZT has received research support from Humanigen, Inc, unrestricted education support from Gilead, ViiV, and Merck (all to the institution). CFK has received research support grants (to the institution) from NIH, CDC, Gilead Sciences and ViiV; VCM has received investigator-initiated research grants (to the institution) and consultation fees (both unrelated to the current work) from Eli Lilly, Bayer, Gilead Sciences and ViiV; AK, CD, DC, OA, GC are employees of, or consultants to, Humanigen, Inc. VMC and FC are third-party agency consultants to Humanigen; CP is a paid consultant to Gilead. ADB is supported by grants from NIAID (grants AI110173 and AI120698) Amfar (#109593) and Mayo Clinic (HH Sheikh Khalifa Bin Zayed Al-Nahyan Named Professorship of Infectious Diseases). ADB is a paid consultant for AbbVie and Flambeau Diagnostics, is a paid member of the DSMB for Corvus Pharmaceuticals, Equilium, and Excision Biotherapeutics, has received fees for speaking for Reach MD, owns equity for scientific advisory work in Zentalis and Nference, and is founder and President of Splissen Therapeutics.

## LIVE-AIR STUDY GROUP

Drs. Meghan Lewis, Linda Sher, Michael Bowdish, Noah Wald-Dickler, Subarna Biswas, Lydia Lam, Khang Vo, Roy Poblete, May M. Lee, Douglass Hutcheon from the University of Southern California (USC) Keck and LAC Medical Centers; Drs. Zelalem Temesgen, and Andrew D. Badley, MD from the Mayo Clinic, Rochester MN; Drs. Charles D. Burger and Claudia R. Libertin from Mayo Clinic, Jacksonville FL; Dr. Jason Baker from Hennepin Healthcare Research Institute Minneapolis, MN; Dr. Victoria Catterson from BioSymetrics, Inc., New York, NY; Dr. William S. Aronstein from CTI, Covington, KY; Drs. Cameron Durrant, Dale Chappell, Omar Ahmed, and Gabrielle Chappell from Humanigen, Inc., Burlingame, CA; Drs. Robert Orenstein and Roberto Patron from Mayo Clinic Arizona; Drs. Vincent C. Marconi, Colleen F. Kelley, John Gharbin, Caitlin Moran, Sheetal Kandiah, Valeria Cantos, Paulina Rebolledo, Carlos del Rio, Jeffrey Lennox, Carmen Polito, Paulina Rebolledo, Anandi Sheth from Emory University Medical Center and Grady Memorial Hospital; Drs. Anup Patel, Homero Paniagua from St. Barnabas Medical Center; Dr. Seife Yohannes from MedStar Washington Hospital Center; Drs. Alpesh Amin, Richard Lee, Miki Watanabe, Lanny Hsieh from the University of California-Irvine Medical Center; Drs. Martin Cearras, Amay Parikh, Jason Sniffen, Wilfred Onyia from AdventHealth Orlando; Drs. Christopher Polk, Michael Boger, Lisa Davidson, Kiran Gajurel, Michael Leonard, Lewis McCurdy, Nestor Quezada, Mindy Sampson, Zainab Shahid, Stephanie Strollo, David Weinrib, Sara Zulfigar from Atrium Health; Drs. Cheryl McDonald, John Hollingsworth, John Burk, Joshua Berg, Daniel Barbaro, Andrew Miller, Lakshmi Sambathkumar, Stuart McDonald, Obinna Okoye from Texas Health Harris Methodist Hospital Fort Worth; Drs. Juan Pulido, Jennifer Fulton, William Gill from Baptist Health Research Institute Jacksonville; Drs. Richard Zuckerman, Lionel Lewis from Dartmouth-Hitchcock Medical Center; Dr. Chaitanya Mandapakala from St. Elizabeth Medical Center; Drs. Matthew Robinson, Brian Metzger from St. David’s Medical Center; Drs. Maqsood Alam, Chrisoula Politis from Mercy Medical; Drs. Anne Frosch, Linh Ngo from Hennepin Healthcare; Drs. Fernando Carvalho Neuenschwander, Estevão Figueiredo, Gualter Cançado, Gustavo Araujo, Lucas Guimarães from Hospital Vera Cruz (NUPEC) in Brazil; Drs. Ricardo Diaz, Natalia Bacellar, Celso Silva, Paulo Ferreira, from Escola Paulista de Medicina (UNIFESP) in Brazil; Dr. Marina Andrade Lima, Caroline Uber Ghisi, Camila Anton, Ricardo Albaneze from Hospital Dia do Pulmãoin Brazil; Dr. Daniel Wagner de Castro Lima Santos, Ana Caroline Iglessias, Marianna Lago, Paula Pietrobom, Maysa Alves from Hospital São Luiz do Jabaquara (IDOR) in Brazil; Drs. Juvencio José Duailibe Furtado, Leopoldo Trevelin, Valeria Telles, Francini Correa from Hospital Heliópolis in Brazil; Drs. Fabiano Ramos, Marina de A. R. Da Silva, Rebeca C. Lacerda Garcia, Ana Elizabeth G. Maldonado, Ana Carolina M. Beheregaray, Ana Maria T. Ortiz from Hospital São Lucas (PUCRS); Drs. Kleber Luz, Eveline Pipolo Milan, Janine Soares de Castro, Matheus José Barbosa Moreira, Renata Bezerra Onofre, Tácito do Nascimento Jácome, Victor Barreto Garcia, Victor Matheus Rolim de Souzafrom Centro de Pesquisas Clínicas de Natal (CPCLIN) in Brazil; Drs. Felipe Dal Pizzol, Cristiane Ritter, Marcelo B. Vinhas from Sociedade Literaria e Caritativa Santo Agostinho (SLCSA) in Brazil; Drs. Adilson Joaquim Westheimer Cavalcante, Julia Minghini, Loni Dorigo, Marina Salgado Miranda from Centro Multidisciplinar de Estudos Clínicos (CEMEC) in Brazil; Drs. Martti Anton Antila, Rebeca Brugnolli, Henrikki Antila from Consultoria Médica e Pesquisa Clínica (CMPC) in Brazil.

## STATEMENTS

### Funding

Humanigen, Inc. the sponsor of LIVE-AIR funded all aspects of the LIVE-AIR study, participated in data collection analysis, interpretation, writing of the manuscript and the decision to submit the manuscript for publication. Humanigen members OA, DC, GC, and CD had access to the raw data.

### Competing interests

All authors have completed the ICMJE uniform disclosure form at www.icmje.org/disclosure-of-interest/ and declare: ZT has received research support from Humanigen, Inc, unrestricted education support from Gilead, ViiV, and Merck (all to the institution); CP is a paid consultant to Gilead; CFK has received research support grants (to the institution) from NIH, CDC, Gilead Sciences and ViiV; VCM has received investigator-initiated research grants (to the institution) and consultation fees (both unrelated to the current work) from Eli Lilly, Bayer, Gilead Sciences and ViiV; CD, DC, OA, AK, and GC are employees of, or consultants to, Humanigen, Inc.; VMC and FC are third-party agency consultants to Humanigen.

### Compliance

The study was conducted in accordance with the Good Clinical Practice guidelines of the International Council for Harmonization E6 and the principles of the Declaration of Helsinki. The protocol was approved by the central/local institutional review board or ethics committee at each site. Participants provided written informed consent.

### Affirmation

The lead author affirms that the manuscript is an honest, accurate, and transparent account of the study. No important aspects of the study have been omitted.

### Data Sharing

Results other than those reported herein will not be shared publicly.

### Provenance and peer review

Not commissioned; externally peer reviewed.

## Notes

### Clinical Trial

NCT04351152

### Funding Statement

This study was funded by Humanigen, Inc. Burlingame, CA

### Author Declarations

The study was conducted in accordance with the Good Clinical Practice guidelines of the International Council for Harmonization E6 and the principles of the Declaration of Helsinki. In the U.S., central IRB (ADVARRA) approval was obtained in addition to local IRB approval at each participating institution, as appropriate. In Brazil, regulatory approval was granted from the Brazilian Health Surveillance Agency (Agencia Nacional de Vigilancia Sanitaria, ANVISA) and approvals from the National Commission of Ethics and Research (Comissao Nacional de Etica em Pesquisa, CONEP) and the Institutional Ethics Committee (Comite de Etica em Pesquisas, CEP) were obtained. All aspects of this company sponsored phase 3 clinical trial (protocol HGEN003 06) were conducted in accordance with the National Health Council (Conselho Nacional de Saude, CNS), Ministry of Health and other applicable regulatory requirements in Brazil. Patients or their legally appointed representative signed written informed consent forms. An independent data and safety monitoring board (DSMB) examined the safety and efficacy of the study medication compared with placebo in addition to standard of care throughout the duration of the study.

## REFERENCES

1. Thwaites RS, Sanchez Sevilla Uruchurtu A, Siggins MK, et al. Inflammatory profiles across the spectrum of disease reveal a distinct role for GM-CSF in severe COVID-19. Sci Immunol. 2021;6(57):eabg9873.

2. Zhou Y, Fu B, Zheng X, et al. Pathogenic T-cells and inflammatory monocytes incite inflammatory storms in severe COVID-19 patients. Natl Sci Rev. 2020;7(6):998–1002.

3. Kox M, Waalders NJB, Kooistra EJ, Gerretsen J, Pickkers P. Cytokine Levels in Critically Ill Patients With COVID-19 and Other Conditions. Jama. 2020;324:1565–1567.

4. Xiong Y, Liu Y, Cao L, et al. Transcriptomic characteristics of bronchoalveolar lavage fluid and peripheral blood mononuclear cells in COVID-19 patients. Emerg Microbes Infect. 2020;9(1):761–770.

5. Weinhold B, Bader A, Poli V, RÜther U. Interleukin-6 is necessary, but not sufficient, for induction of the human C-reactive protein gene in vivo. Biochemical Journal. 1997;325(3):617–621.

6. Zhang D, Sun M, Samols D, Kushner I. STAT3 participates in transcriptional activation of the C-reactive protein gene by interleukin-6. J Biol Chem. 1996;271(16):9503–9509.

7. Lavillegrand JR, Garnier M, Spaeth A, et al. Elevated plasma IL-6 and CRP levels are associated with adverse clinical outcomes and death in critically ill SARS-CoV-2 patients: inflammatory response of SARS-CoV-2 patients. Ann Intensive Care. 2021;11(1):9.

8. Petrilli CM, Jones SA, Yang J, et al. Factors associated with hospital admission and critical illness among 5279 people with coronavirus disease 2019 in New York City: prospective cohort study. Bmj. 2020;369:m1966.

9. Hodges G, Pallisgaard J, Schjerning Olsen AM, et al. Association between biomarkers and COVID-19 severity and mortality: a nationwide Danish cohort study. BMJ open. 2020;10(12):e041295.

10. Manson JJ, Crooks C, Naja M, et al. COVID-19-associated hyperinflammation and escalation of patient care: a retrospective longitudinal cohort study. Lancet Rheumatol. 2020;2(10):e594–e602.

11. Keller MJ, Kitsis EA, Arora S, et al. Effect of Systemic Glucocorticoids on Mortality or Mechanical Ventilation in Patients With COVID-19. J Hosp Med. 2020;15(8):489–493.

12. Placais L, Richier Q, Noel N, Lacombe K, Mariette X, Hermine O. Immune interventions in COVID-19: a matter of time? Mucosal Immunol. 2021.

13. Temesgen Z, Burger CD, Baker J, et al. Lenzilumab Efficacy and Safety in Newly Hospitalized Covid-19 Subjects: Results from the Live-Air Phase 3 Randomized Double-Blind Placebo-Controlled Trial. Lancet Respir Med. 2021; DOI:https://doi.org/10.1016/S2213-2600(21)00494-X.

14. Keddie S, Ziff O, Chou MKL, et al. Laboratory biomarkers associated with COVID-19 severity and management. Clin Immunol. 2020;221:108614.

15. Beigel JH, Tomashek KM, Dodd LE, et al. Remdesivir for the Treatment of COVID-19 - Final Report. N Engl J Med. 2020;383(19):1813–1826.

16. Booz GW, Altara R, Eid AH, et al. Macrophage responses associated with COVID-19: A pharmacological perspective. Eur J Pharmacol. 2020;887:173547.

17. Blot M, Bour JB, Quenot JP, et al. The dysregulated innate immune response in severe COVID-19 pneumonia that could drive poorer outcome. J Transl Med. 2020;18(1):457.

18. Szalai A, van Ginkel F, Dalrymple S, Murray R, McGhee J, Volanakis J. Testosterone and IL-6 Requirements for Human C-Reactive Protein Gene Expression in Transgenic Mice. The Journal of Immunology. 1998;160(11):5294–5299.

19. RECOVER Collaborative Group. Tocilizumab in patients admitted to hospital with COVID-19 (RECOVERY): a randomised, controlled, open-label, platform trial. The Lancet. 2021;397(10285):1637–1645.

20. REMAP-CAP Investigators, Gordon AC, Mouncey PR, et al. Interleukin-6 Receptor Antagonists in Critically Ill Patients with Covid-19. N Engl J Med. 2021;384(16):1491–1502.

21. Martinez-Sanz J, Muriel A, Ron R, et al. Effects of tocilizumab on mortality in hospitalized patients with COVID-19: a multicentre cohort study. Clinical microbiology and infection : the official publication of the European Society of Clinical Microbiology and Infectious Diseases. 2021;27(2):238–243.

22. Mariette X, Hermine O, Tharaux P, et al. Effectiveness of Tocilizumab in Patients Hospitalized With COVID-19: A Follow-up of the CORIMUNO-TOCI-1 Randomized Clinical Trial. JAMA Internal Medicine. 2021.

23. Health NIo. COVID-19 Treatment Guidelines: Interleukin-6 Inhibitors. https://www.covid19treatmentguidelines.nih.gov/therapies/immunomodulators/interleukin-6-inhibitors/ Accessed August 18, 2021. Published 2021. Updated April 21, 2021. Accessed August 18, 2021.

